# The establishment of the first ex vivo whole organ model for human liver neoplasms

**DOI:** 10.1101/2020.07.05.20146290

**Authors:** Qiang Zhao, Jingjing Li, Caihui Zhu, Honghui Chen, Yihao Ma, Weixin Luo, Rongxing Xie, Yixi Zhang, Xiaobo Wang, Linhe Wang, Zhiheng Zhang, Zhiyong Guo, Xiaoshun He

## Abstract

**Background:** The incidence of liver neoplasms is on the leading rise worldwide due to lacking exact research model. Accordingly, the resected diseased liver within cancer during liver transplantation was the appropriated model, therefore the aim of this study was to establish the first ex vivo whole organ model for liver neoplasms by using normothermic perfusion system named Life-X system.

**Materials and Methods:** Four diseased livers within cancer resected during liver transplantation were collected for research. The common hepatic artery and portal vein of the ex vivo liver were connected to the Life-X perfusion device that circulated Life-X perfusate providing continuous oxygen and nutrient supply. The flow and pressure of the perfusate was recorded and blood gas analysis was examined to analyze the function of the diseased liver. Liver tissues after perfusion were collected for histological analysis.

**Results:** Experiments showed that the artery and portal vein flow were stable 1h after perfusion and were kept within the physiological range. The results of blood gas analysis demonstrated restoration and maintenance of metabolism. Moreover, the bile production of diseased Case 4 liver represented its vivid functionality during the entire 47h of perfusion. Histology analysis shows little liver injury after the perfusion.

**Conclusions:** Therefore, we have established a powerful tool to research liver neoplasms in vitro through Life-X perfusion system.

## Background

Nowadays, there are more than 800,000 new cases of liver neoplasms and 780000 deaths annually, making it the sixth prevalent and the fourth fatal cancer worldwide.^1^ In China, almost 7 million people are suffering from liver cirrhosis, leading to 460,000 new liver cancer cases annually.^2^ This public human health threat has become much worse due to a shortage of effective curative interventions, which are strapped for lacking exact research model to explore its pathogenic mechanism as well as to test the effect of therapeutic drugs.^3^

Pre-clinical research models have consisted of human liver cancer cell lines and animal models.^3,4^ Vast of explorations have been made to recognize the character and pathophysiology of liver neoplasms in these cell or animal based models, however, few were translated to bedside because of their failure to mimic highly heterogeneity or the internal environment of tumor. Thence, new and improved research models are in urgent need that can be used not only to study the pathophysiological processes of liver cancer, but also in drug discovery.

Normothermic machine perfusion (NMP) is an innovative technique in organ transplantation by continuously supplying nutrients and oxygen at physiologic temperature to maintain organ activity including marginal donor livers.^5^ And NMP has been proven to reverse energy deficits and inhibit inflammation to maintain the stability of internal environment, thus it could protect donor organ from ischemia and reperfusion injury and improve organ quality prior to transplantation.^6^ However, there is little research about diseased liver within cancer under normothermic machine perfusion.

Therefore, we postulate that, under appropriate culture conditions, certain molecular and cellular functions of the resected diseased liver could be maintained for a long time that can be used as an ideal research platform for liver cancer. To test this hypothesis, we developed a surgical procedure, perfusate, and custom pulsatile-perfusion device that can restore and maintain microcirculation and cellular viability in human liver cancer under ex vivo normothermic conditions (37 °C). This system is herein referred to as Life-X system. Here, we describe the establishment of an ex vivo whole organ model for studying human liver neoplasms. With this new model, we were able to study liver neoplasms in near physiological conditions and test therapies response in the near future.

## Materials and Methods

### Overview of Life-X technology

The technology consists of a perfusion system that circulates Life-X perfusate (Table 1) (Fig. 1A). The system is amenable to any custom waveform within 10-180 mm Hg as well as temperatures of 37-42°C. Moreover, the platform supports organ homeostasis through gas and fluid infusion mechanisms. We also developed a surgical procedure for isolating the diseased liver and its vascular supply during liver resection. The lower common bile duct was transected firstly, followed by intermediate resection of the common hepatic artery (HA) and portal vein (PV). The liver was then retrieved by resection of the suprahepatic and infrahepatic vena cava. An immediate cold flush through both hepatic artery and the portal vein was carried out with 1500 mL normal saline and 37500U heparin at 4 °C, to wash out the residue blood of the collected livers. The common hepatic artery and portal vein were intubated and connected to the Life-X perfusion device (Fig. 1B) and ex vivo circulation was maintained for a length of perfusion of 11 h. The study protocol was approved by the Ethical Committee of The First Affiliated Hospital of Sun Yat-sen University, and informed consent was obtained from the participant.

**Table 1.** Components of the Life-X perfusate solution.

|  |  |
| --- | --- |
| Succinylated gelatinor | 1.2 L |
| Crossed-matched leucocyte-depleted washed red cells | 1.2 L |
| Metronidazole | 0.5 g |
| Cefoperazone sodium and sulbactam sodium | 1.5 g |
| 5% sodium bicarbonate | 140 ml |
| Heparin | 20000U |
| 10% calcium gluconate | 35 mL |
| 25% magnesium sulphate | 3 ml |
| Methylprednisolone sodium succinate | 500 mg |

**Figure 1:**
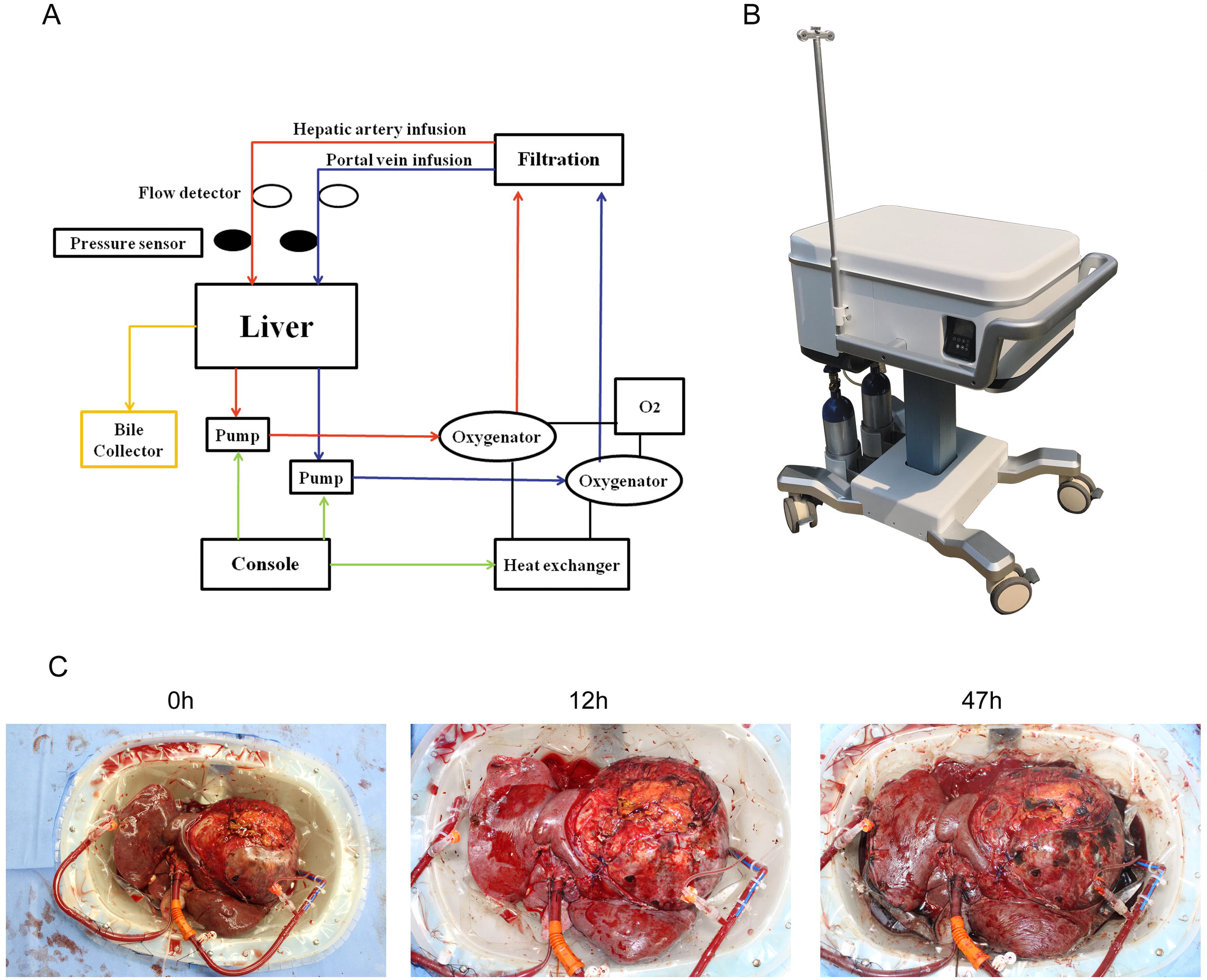
The Life-X perfusion system was presented. (A) It consists of an organ chamber, a console, two roller pump, a heat exchanger, two oxygenator, two pressure sensors, two flow detectors and a bile collector. (B)The macroscopic picture of Life-X perfusion system. (C)The macroscopic picture of Case 4 liver during 47 hours of perfusion.

### Preservation of the liver function

Under Life-X perfused conditions, oxygen was administered at 200 mL/min, oxygen partial pressure was maintained at 250–500 mmHg, while carbon dioxide partial pressure kept at 30–50 mmHg. The HA flow was maintained at 90–240 mL/min with a perfusion pressure of 50–70 mmHg, and the PV flow was maintained at 0.3–1.4 L/min with a pressure of 8–13 mmHg. During the perfusion, 5% glucose, insulin and compound amino acid were continuously infused into the circuit. The blood gas analysis of perfusate in all the cases as well as bile production and anslysis in Case 4 liver were performed in different time point.

### Histological analysis

For histological analysis, liver tissues of all 4 cases were collected to Paraffin-embedded slides at the end of perfusion. And those slides were deparaffinized with xylene and dehydrated with graded alcohols, then hematoxylin, eosin (HE) staining was performed according to the manufacturer’s instructions.

### Statistical analysis

Statistical analysis was performed with the SPSS software (19.0; SPSS, Inc., Chicago, IL). Values are expressed as mean ± standard deviation (SD). The Student’s t test was used for comparisons between groups. P < 0.05 was considered statistically significant.

## Results

### Stable vascular perfusion by Life-X

Four patients aged from 37-75 year-old were diagnosed with liver neoplasms, with or without hepatitis B (HBV) infection and underwent liver transplantation in our hospital in 2019 (Table 2).

**Table 2.** Characteristics of the patients.

| Characteristics | Case 1 | Case 2 | Case 3 | Case 4 |
| --- | --- | --- | --- | --- |
| Sex | Male | Male | Male | Male |
| Age (year) | 56 | 65 | 37 | 75 |
| Height (cm) | 171 | 170 | 168 | 168 |
| Weight (kg) | 67 | 64 | 100 | 67 |
| Blood type (Rh) | O(+) | B(+) | A(+) | O(+) |
| Diagnosis | HCC | HCC | HCC | HCC |
| HBsAg | negative | positive | negative | negative |
| HBV DNA (IU/mL) | <100 | <100 | <100 | <100 |
| ALT (U/L) | 36 | 72 | 146 | 19 |
| AST (U/L) | 79 | 59 | 401 | 31 |
| ALP (U/L) | 151 | 156 | 61 | 98 |
| GGT (U/L) | 31 | 123 | 61 | 148 |
| LDH (U/L) | 356 | 167 | 412 | 226 |
| Total Bilirubin (umol/L) | 65.6 | 10.5 | 43.4 | 17.8 |
| Child-Pugh grade | A | A | A | A |
| Perfusion time (h) | 10 | 11 | 18 | 47 |

During the entire perfusion, the Life-X system was stable, and no technical problems were observed (Fig. 1C). The mean hepatic artery flow was kept stable 1h after initiation of the perfusion range from 100-230 ml/min (Fig. 2B) with mean perfusion pressure at 52 to 60 mmHg (Fig. 2A). The hepatic artery resistance index was gradually dropped from 0.6 to 0.3 (Fig. 2C). The portal vein flow was maintained stable at 0.5-1.3 L/min 1h after perfusion (Fig. 2E) with the pressure stabilizing at 8-13 mmHg (Fig. 2D). And the level of portal vein resistance index was dropped from 27 to nearly 8 (Fig. 2F). These findings suggested that the microvasculature was patent and maintained flow under Life-X perfusion and both the artery and portal vein flow were kept at the physiological state.

**Figure 2:**
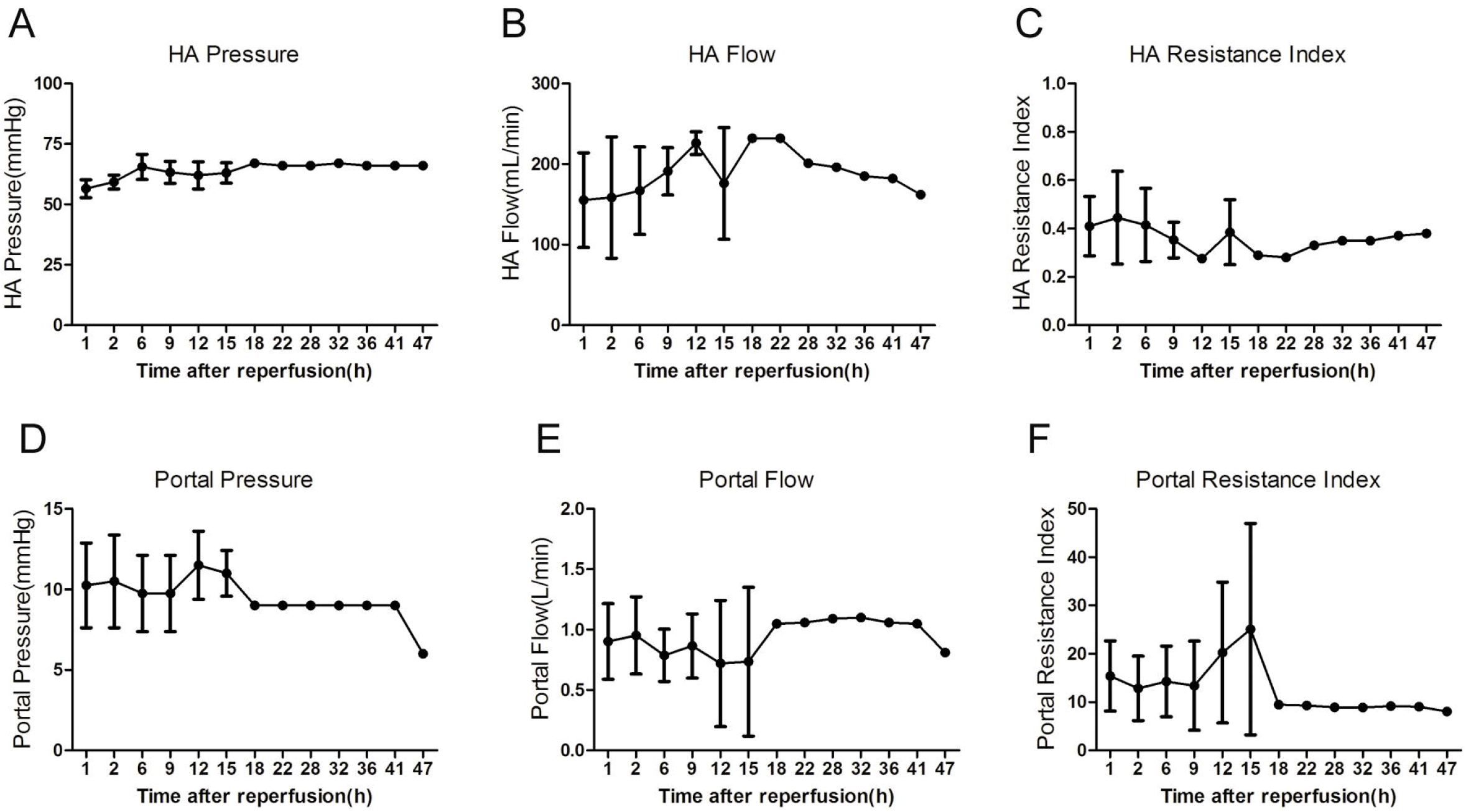
The blood flow and pressure of all four livers during the perfusion. (A) The pressure of hepatic artery. (B) The flow of hepatic artery. (C) The resistance index of hepatic artery. (D) The pressure of portal vein. (E) The flow of portal vein. (F) The resistance index of portal vein.

### Preservation of the diseased liver function

The arterial carbon dioxide partial pressure (pCO_2_) of all four livers was kept stable range from 15 mmHg to 75 mmHg (Fig. 3A), while the oxygen partial pressure (pO_2_) was fluctuated between 90-340 mmHg (Fig. 3B) indicating a normal oxygenated condition conducted by Life-X. And the mean perfusate glucose was fluctuated between 27.8 to 3.4 mmol/L (Fig. 3C), which means a significantly glucose consumption during perfusion. Also the potential of hydrogen (pH) value was maintained stable at the normal range (Fig. 3D), while the mean level of perfusate lactate was retain lower than 5 mmol/L during most of the perfusion time (Fig. 3E). Persistent bile production during perfusion was observed in Case 4 liver at 1.0-2.5 ml per hour (Fig. 3F), while the level of pCO_2_, pO_2_, pH and glucose in the bile was stable at the normal range (Fig. 3G-J). Overall, these data demonstrated restoration and maintenance of metabolism and bile production of the diseased liver for as long as 47 h ex vivo by Life-X system.

**Figure 3:**
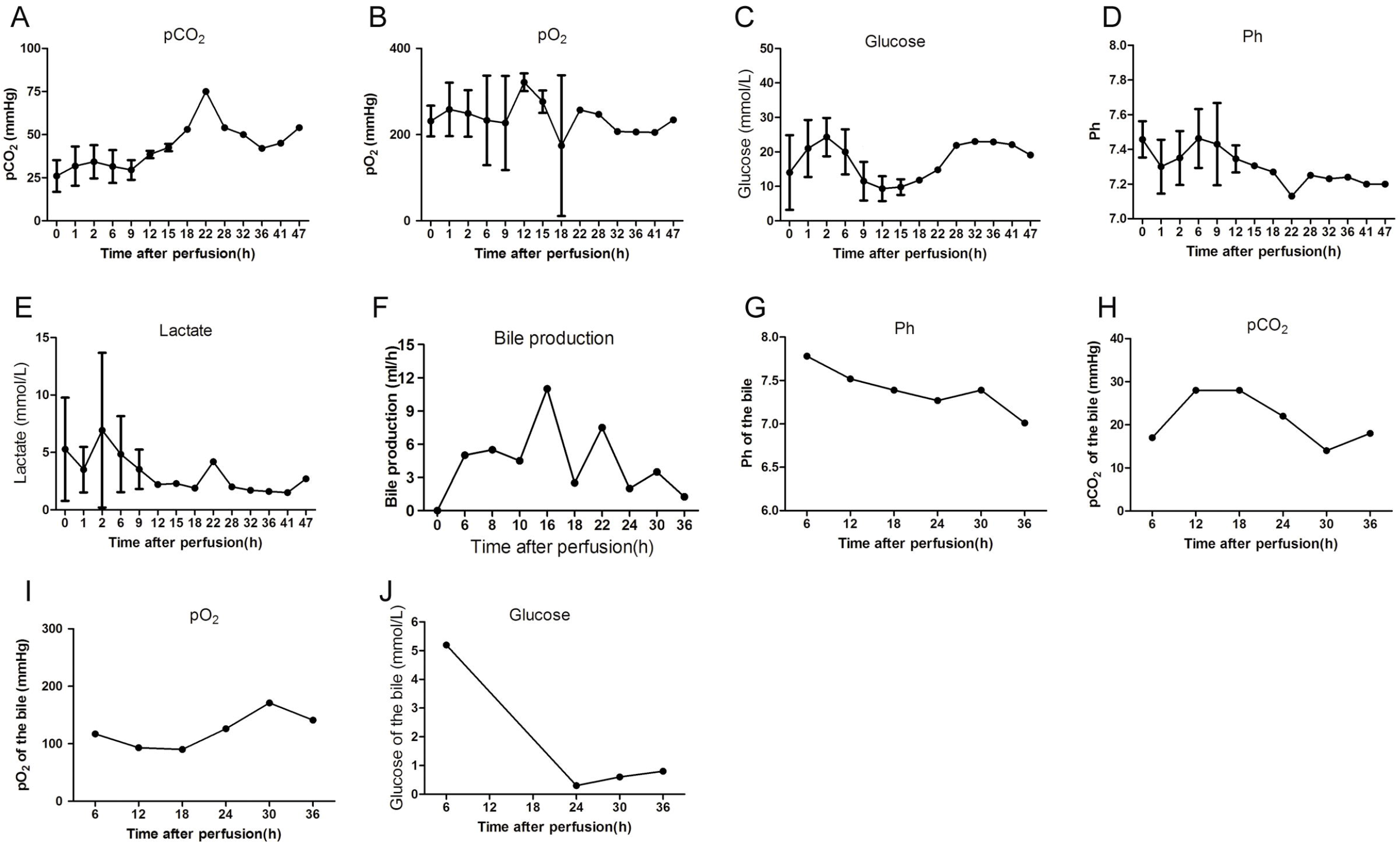
Blood gas analysis of all four livers as well as bile production and analysis of Case 4 liver. (A) The arterial carbon dioxide partial pressure (pCO_2_) of all four livers. (B) The oxygen partial pressure (pO_2_) of all four livers. (C) The level of perfusate glucose of all four livers. (D) The potential of hydrogen (pH) value of all four livers. (E) The level of perfusate lactate of all four livers. (F) Bile production of Case 4 liver. (G) The potential of hydrogen (pH) value of the bile of Case 4 liver. (H) The carbon dioxide partial pressure (pCO_2_) in the bile of Case 4 liver. (I) The oxygen partial pressure (pO_2_) in the bile of Case 4 liver. (J) The level of glucose in the bile of Case 4 liver.

### Pathological changes

The liver graft appeared no obvious change and homogeneously perfused. HE staining analysis showed no changed parenchyma, preserved sinusoidal architecture and viable hepatocytes after the perfusion (Fig. 4 A-D). No bacterial growth was detected in the perfusate sampled at the beginning and end of perfusion. Perfusion was discontinued as determined based on the availability of the research team to continuously monitor the organ. Overall, the liver tissue integrity was preserved, and no further damage was observed during Life-X support.

**Figure 4:**
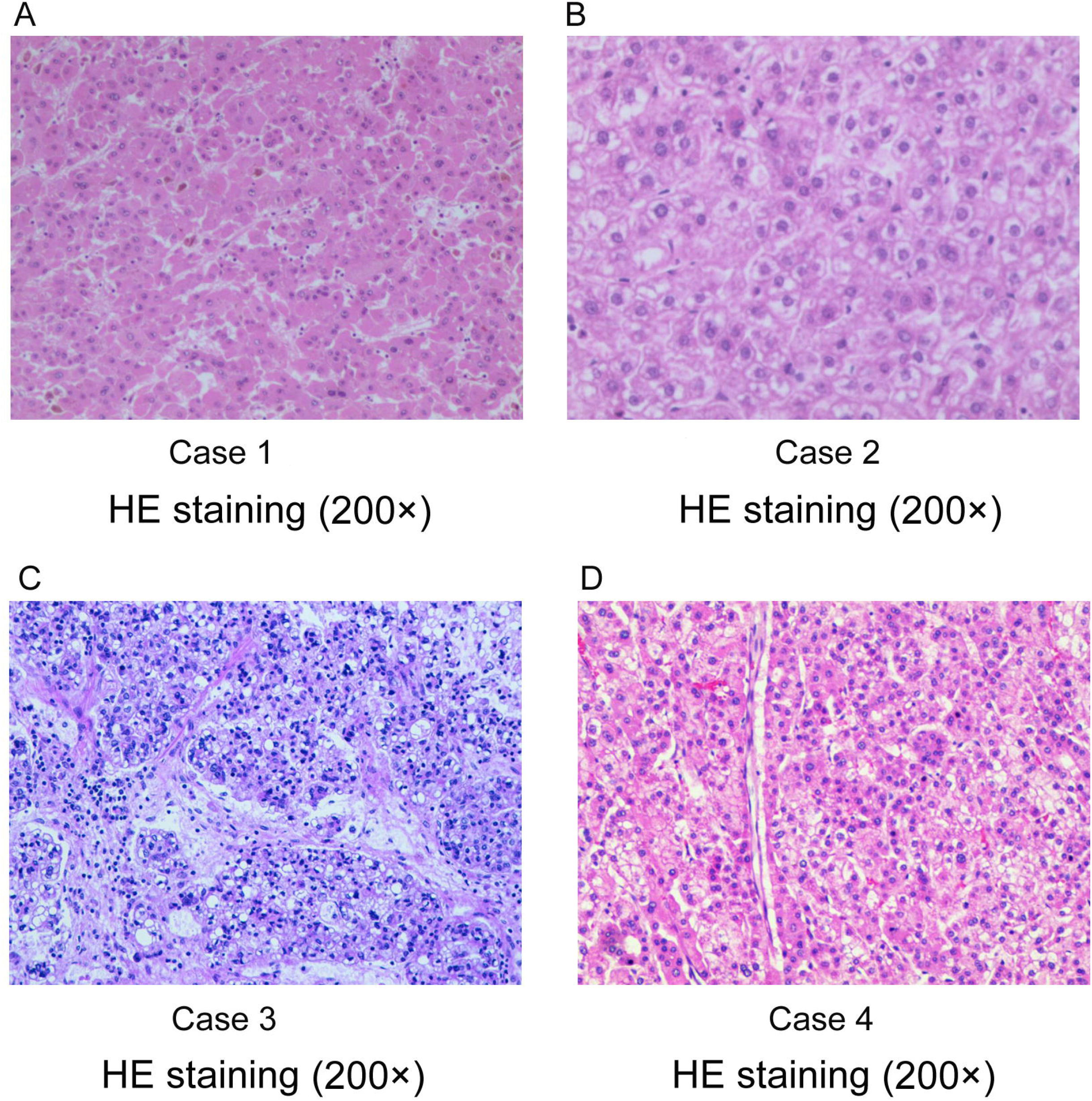
Histology analysis of liver after the normothermic machine perfusion of all four cases. (A-D) Hematoxylin, eosin (HE) staining (200_×_) showed little changes after the perfusion.

## Discussion

The Life-X perfusion system has provided a beneficial and clinical feasible platform for donor livers preservation, increasing the number of donor livers and improving the marginal organ function, under which we have invented a new procedure ischemia-free liver procurement (IFLP) to reduce the hepatocyte and biliary epithelium injury.^7,8^ Recently, we have firstly proposed a new concept ischemia-free organ transplantation (IFOT) that implanting a liver graft with continuous nutrient supply under perfusion preservation had completely avoided ischemia and reperfusion injury (IRI). ^8,9^

Studies reveal that the existing human cancer-derived cell lines have served as an essential platform for exploring mechanism of cell action and genomic signature of primary tumor tissue, as well as efficiently manifesting drugs screening in the early stages of pre-clinical drug development.^10^ However, cell-based studies often disregard the interactions of cell to cell or cell to matrix, and their natural system components in 2D culture systems. Other limitations are exemplified by frequently mutations in essential genes like TP53 in such cell lines, resulting in hardly representing patient-specific signatures.^11^ Promising models like 3D culture systems termed “organoids” derived from human primary liver cancer tissues could faithfully model its genetic complexity, and self-renew and differentiate in vitro.^12^ Its reproduction makes organoids a valuable resource in identifying essential roles in liver neoplasms and a potential personalized-medicine approach in screening novel drugs. Lacking stromal components and immune system, organoids could hardly reproduce tumor heterogeneity, tumor immune microenvironment and tumor vascular network in vitro. ^13^

Unfortunately, due to species tropism and the difficulties in culturing human hepatocytes, there were limited systems for studying viral hepatitis, a leading cause of hepatocellular carcinoma (HCC), particularly in Asian countries.^14^ Our interesting finding was that a whole liver within cancer could maintain its viability during the entire perfusion. Continued lactate clearance and bile production represented that the function of liver was well preserved. Histological analysis showed that no further injury was detected during perfusion. These results reflected that Life-X perfusion system could apply to be a reliable platform to support investigation for human liver neoplasms.

Based on that Life-X system provides the opportunities to monitor organ function, our system might be extendedly applied to other dysfunctional livers such as liver cirrhosis, liver failure and portal vein hypertension, or other human organs such as kidey, lung and heart. We have explored out the maintenance condition of kideys from circulatory death (DCD) donors and expanded criteria donors (ECD),^15^ which could build up our personalized isolated organ research platform. And we have attempted to combined liver-kidney perfusion to protect livers from warm ischemia injury in pig experiments.^16^

The controllable perfusion fluid content offers possibility to combine other therapies with Life-X. Several attempts have been made to add pharmacologic treatments during perfusion. Evidence showed that drug delivery like hydrogen sulphide with machine perfusion has been proven effective in reducing pig’s renal ischaemia-reperfusion injury.^17^ Cell therapy like mesenchymal stromal cells could be administered during perfusion before kidney transplantation in a humanized mouse model to reduce graft rejections as injured kidney cells could be directly targeted^18^.

In vitro liver neoplasms model representing individual patients will facilitate the development of personalized medicine. It is promising that combine large-dose chemotherapy drugs with perfusion to make individualized treatment for liver cancer patients. Large-scale drugs screening will become much easier through our systems.

This is an important finding as establishing a novel model for liver neoplasms research and could provide an opportunity for pharmacologic, immunologic and genetic interventions. Our technology requires further development, optimization, and implementation, including studies with longer perfusion times. This new research model may have broader applications than those described herein, and could potentially help to bridge the gap between basic medicine science and clinical research.

In summary, we have established a powerful tool to research liver neoplasms in vitro through Life-X system, and our system will eventually developed into an inspiring research models for diseased human organs.

## Data Availability

All data are contained in the manuscript.

## Funding

Our research was supported by the National Natural Science Foundation of China (81970564, 81471583, 81570587 and 81700557), the Guangdong Provincial Key Laboratory Construction Projection on Organ Donation and Transplant Immunology (2013A061401007 and 2017B030314018), Guangdong Provincial Natural Science Funds for Major Basic Science Culture Project (2015A030308010), Guangdong Provincial Natural Science Funds for Distinguished Young Scholars (2015A030306025), Special Support Program for Training High Level Talents in Guangdong Province (2015TQ01R168), Science and Technology Program of Guangzhou (201704020150), the Natural Science Foundations of Guangdong province (2016A030310141 and 2020A1515010091) and Young Teachers Training Project of Sun Yat-sen University (K0401068).

## Declaration of Conflicting Interests

The authors declared that there is no conflict of interest.

## Abbreviations

ALP: alkaline phosphatase
ALT: alanine transaminase
AST: aspartate transaminase
DCD: donation after cardiac death
ECD: expanded criteria donors
GGT: gamma-glutamyl transferase
HA: hepatic artery
HBV: Hepatitis B virus
HBsAg: Hepatitis B surface antigen
HBV DNA: Hepatitis B virus deoxyribonucleic Acid
HCC: hepatocellular carcinoma
HCO_3_-: bicarbonate
HE: hematoxylin and eosin
IFLT: ischemia-free liver transplantation
IFOT: ischemia-free organ transplantation
IRI: ischemia and reperfusion injury
LDH: lactate dehydrogenase
PV: portal vein
pCO_2_: arterial carbon dioxide partial pressure
pO_2_: oxygen partial pressure.

## Notes

### Competing Interest Statement

The authors have declared no competing interest.

### Author Declarations

The study protocol was approved by the Ethical Committee of The First Affiliated Hospital of Sun Yat-sen University, and informed consent was obtained from the participant.

